# The Role of Home Medication Storage Location in Increasing Medication Adherence for Older Adults

**DOI:** 10.1101/2022.07.21.22277904

**Authors:** Lisa Gualtieri, Eden Shaveet, Brandon Estime, Avi Patel

**Affiliations:** Tufts University School of Medicine, Department of Public Health and Community Medicine, Boston, MA, 02111, USA

**Keywords:** medication adherence, older adults, digital health, medication management

## Abstract

**Background:** Over 50% of U.S. adults do not take their prescriptions as prescribed, which is responsible for 33% to 69% of hospital admissions and 125,000 deaths annually. Given the higher prevalence of prescription drug use among older adult populations, increasing medication adherence is of particular importance with this age group. Two speculated facilitators of medication adherence are home medication storage location and the use of digital health devices.

**Objective:** Our objective was to use survey data to investigate the associations between medication storage location and medication adherence among adults 40 years and older. Additionally, we aimed to report preliminary findings about the associations between use of digital health devices and medication adherence in this same population.

**Methods:** We conducted primary analysis of data sampled from a home medication management survey deployed in November 2021 (n=580). Exploratory analyses were conducted by way of chi^2^ tests and creation of bivariate logistic regression models.

**Results:** The most heavily used storage locations were nightstand drawers (27%), kitchen cabinets (25%), and atop bedroom nightstands (23%). Certain medication storage locations were associated with greater medication adherence. Several storage locations were significantly associated with decreased odds of having ever forgotten to take a medication, including kitchen drawers, in refrigerators, atop bedroom nightstands, in nightstand drawers, and backpacks, purses, or bags. Two home medication storage locations were significantly associated with increased odds of having ever forgotten to intake a medication: kitchen cabinets and bathroom vanities. Further, most (94%) survey respondents indicated they would be receptive to guidance about where to store their medications.

**Conclusions:** Given that all patients need to select a location to store their prescription medication in their homes and that some storage locations are associated with adherence, we believe that an intervention to optimize storage selection may lead to increased adherence. The impact of digital health device usage paired with optimized home medication storage location on medication adherence in older adult populations remains unknown but is worthy of further investigation. Further, we plan to investigate how new device designs can incorporate specific storage locations and contextual cues related to location to promote adherence more effectively.

## 1 Introduction

### 1.1 Definition and Importance

Medication adherence, the extent to which a patient follows a stipulatory prescription treatment plan, is crucial to the success of patient care and is indispensable for reaching clinical goals (1). However, only 50% of people adhere to medication guidelines (2), posing substantial risk to patient health and safety. Medication nonadherence is responsible for as many as 33% to 69% of hospital admissions and 125,000 deaths annually (3). Other consequences of medication nonadherence include waste of medication, disease progression, and an overall lower quality of life (4). There is a rising trend in medication nonadherence across all age, sex, and racial groups in the US (5), which is particularly concerning for older adults as the likelihood to be prescribed long term medications increases with age (6, 7).

Medication nonadherence, while well-studied, continues to be a vexing issue for clinicians and researchers (2, 8). In 2003, the World Health Organization (WHO) indicated that improved medication adherence interventions may have greater impact on population health than specific medical treatments (9). Known barriers to medication adherence include out-of-pocket costs of medications, physical difficulties obtaining a medication supply, health literacy, and difficulty following intake guidance (10, 11). Patients facing multiple, co-occurring barriers are even less likely to adhere to prescribed medication (12).

### 1.2 Home Medication Management

Given the importance of contextual cues and mental associations in developing routines, medication storage locations may be a vital component of improving medication adherence. In the process of habit formation, settings that are associated with certain behaviors influence an individual’s actions to deviate from conscious motivation (13). External contextual cues in a person’s environment trigger an automatic response that promotes habit formation (14). Previous work exploring strategies for medication adherence found that reliance on multiple cues may have a beneficial impact on adherence to medication regimens (15). Locations where medications are frequently stored may have the capacity for a plethora of contextual cues.

Understanding a patient’s home medication management routine, including home medication storage location selection, may be needed for more effective medication management in general, and for the locating of devices that dispense or remind in particular. For example, devices that rely on auditory or visual cues need to be in locations where patients can hear or see the alerts and notifications. While home medication management is well-studied in relation to patient safety (16), less is known about the role of home medication storage location in medication adherence.

### 1.3 Patients and Locations of Interest

Behavior change theories such as the Rubicon model of action phases suggest that the level of intent in improving medication adherence should be considered when developing medication adherence interventions (17). According to the this model, interventions are most effective when an individual intends to be adherent to their medication but only needs additional support and guidance to do so (18). As a result of exploring this theory, our primary interest is in patients who either do not face or have overcome well-documented barriers to medication adherence, such as cost and access, and intend to be adherent, yet still struggle to achieve adherence (18).

Our population of interest is older adults as this demographic is more likely to take one or more prescriptions compared to other age groups (6, 7). For some health conditions such as hypertension and diabetes mellitus, a patient’s older age may also impact their level of adherence (19, 20).

Our location of interest is in patients’ homes as more medications are taken at home than in hospitals and clinics combined (1, 21). Our research focuses on understanding the barriers to medication adherence in the home in order to design aids that increase adherent behavior. We explore this by first understanding where patients store their medications, and then by investigating how these locations are selected and if it is the locations themselves or the determinants of selection that correlate with adherence. Finally, we examine factors related to home medication management including the use of digital and non-digital aids to help with adherence.

### 1.4 Home Medication Storage Locations

Prior research into home medication storage has examined storage conditions with respect to safety, climatic conditions, and routines; however, the role of home storage location’s impact in medication adherence is understudied in literature. A study of medication storage conditions found that, of 170 participants aged 65 and older, 76% complied with drug product label recommendations for temperature, light, and humidity (22). A study on medication disposal found that 81.5% of respondents in 445 telephone interviews had prescription medications and almost all the respondents indicated that there were excess and leftover medications in their homes (23). Another study on safe medication storage surveyed 1,074 people aged 50-80 with grandchildren aged 0-17 (24). Their findings indicated that 89% of respondents had prescription medications in their homes and 84% reported that they kept their medications in the same place they typically store them when their grandchildren visited (24).

MedlinePlus, an online information service produced by the United States National Library of Medicine, suggests storing medications in the dresser drawer, kitchen cabinet, storage box, shelf, or closet (25). Although they do not reference medication storage location in regard to medication adherence, they state that location plays a role in medication effectiveness and safety. With 418 million users in 2021 (26), MedlinePlus could influence medication storage decisions.

A few studies have addressed the role of strategies, routines, and habit formation in establishing a successful medication regimen (15, 27). One study found that older adults employ both internal (e.g., use of mental associations) and external (e.g., use of physical objects and/or locations) strategies to remember to take their medications (15). Another study considered the use of contextual cues as an aid to develop a new behavior, reporting that patients who store their medications in locations that are conducive to their routines were more likely to be adherent to their medication regimens (28). Behavioral interventions that provided counseling on adherence strategies considered the storage of medications only as a cue to remind individuals to maintain adherence (29, 30), yet their effect was modest; survey respondents who used cues that were visible throughout the day were unable to remember whether they had taken their medication (28).

Several efforts have been dedicated to the development and use of devices to improve medication adherence, including digital health devices and apps that are designed to dispense medication and/or remind users that it is time to take medication (28). However, simple, low-cost devices have been investigated but have yet to produce clinically impactful outcomes (31). There are an increasing number of devices aimed at improving adherence, yet sub-optimal placement of these devices in the home may lead to less efficacious changes in adherence.

No study has evaluated the relationship between medication storage locations and adherence. Furthermore, no study has evaluated the relationship between storage location and device use. Given these gaps in the literature, we designed and deployed a survey to learn more about medication management in the home.

## 2 Methods

### 2.1 Study Sample

Data were collected via deployment of the Home Medication Management Survey, designed by members of the Digital Health Research Group at Tufts University, and fielded between November 18 and December 14, 2021. The survey was deployed in English via Google Forms, an online, cloud-native survey-development platform whose files are encrypted in transit and at rest (32). Participants were recruited via informational posts on social media platforms, including Twitter, Facebook, Instagram, and LinkedIn, and via the Osher Lifelong Learning Institute at Tufts University mailing list, which reaches an audience of around 2,000 older adults primarily in the Greater Boston area. Eligible participants for the survey were 18 years of age or older with access to an internet-enabled device. Upon completion of the survey, participants could elect to enter a drawing for one of five Amazon gift cards valued at $25. All study protocols were reviewed and approved by the Tufts University Health Sciences Institutional Review Board in Boston, MA.

### 2.2 Procedures

A total of 1,966 survey responses were recorded at the close of the survey, and 1,673 (85%) of responses were deemed to be valid after excluding responses on suspect of fraudulence. Dropped responses met any of the following exclusion criteria: responses were not in English (49 responses dropped); a consecutive set of identical responses were posted at the same time (132 responses dropped); an abnormal email address was provided for the drawing that included a long string of numbers suspected to have been generated by a bot (19 responses dropped); or responses included suspicious identical open-text responses within the same day (83 cases dropped).

Of the 1,673 remaining eligible responses, our study sample for this paper was respondents indicating an age of 40 years or older (n=580). We selected this age range to adhere with the CDC’s inclusion of 40-79 years in their report of prescription drug use (33).

### 2.3 Measures

Survey questions were designed to learn about respondents’ experiences with medication management in the home. Questions included ones about use of aids to adherence, perceived importance of adherence, self-reported adherence, as well as demographic questions.

#### 2.3.1 Demographic Variables

Demographic items included in our analysis were age in years by decade (40-49, 50-59, 60-69, 70-79, 80-89, 90+), race and ethnicity (multi-select categorical item), sex, and highest level of education completed. *Table 1: Sample Characteristics* displays the demographic distributions.

**Table 1:** Sample Characteristics (N = 580)

| Characteristic | n (%) |
| --- | --- |
| **Age in years** |  |
| 40 - 49 | 244 (42) |
| 50 – 59 | 127 (22) |
| 60 - 69 | 82 (14) |
| 70 - 79 | 84 (14) |
| ≥ 80 | 43 (7) |
| **Race/Ethnicity\*** |  |
| White | 439 (76) |
| American Indian or Alaskan Native | 70 (12) |
| Black or African American | 41 (7) |
| Asian | 27 (5) |
| Hispanic or Latino | 27 (5) |
| Native Hawaiian or Other Pacific Islander | 12 (2) |
| **Sex\*\*** |  |
| Female | 345 (59) |
| Male | 233 (40) |
| **Highest Education Completed** |  |
| < High school diploma or equivalency | 17 (3) |
| High school diploma or equivalency | 45 (8) |
| Some college | 82 (14) |
| Associate's degree | 47 (8) |
| Bachelor's degree | 160 (28) |
| Master's or professional degree | 183 (32) |
| Doctoral degree | 46 (8) |

\* Responses non-mutually exclusive; <1% identified with an unlisted race or ethnicity
\*\* <1% identified with an unlisted sex

#### 2.3.2 Digital and Non-digital Medication Reminders

Our measures indicating use of digital and non-digital medication reminder methods were derived from two survey items from which respondents could select all that applied: non-digital methods (“Written notes,” “Post-it notes,” “Calendar,” “Chart,” “Pillbox”) and digital methods (“Smartphone app,” “Smartphone alarm,” “Siri,” “Alexa,” “Smartwatch,” “Electronic pill dispenser,” “GlowCap or attachment to pill bottle”).

#### 2.3.3 Perceived Importance of Medication Adherence

Perceived importance of medication adherence was measured on a five-point Likert scale ranging from “not at all important” to “very important” in response to a survey item asking, “How important is it for you to take your medication as prescribed?”

#### 2.3.4 Medication Adherent Behavior

Our variables assessing medication adherence were derived from questions which asked about forgetting to take a medication as a proxy for adherence (34). Our variables were dichotomous (“yes”, “no”) in response to survey items asking, “Have you ever forgotten to take a medication?” and “In the past two weeks, have you forgotten to take a medication?”

#### 2.3.5 Medication Storage Location

Our variable for storage locations of medications taken regularly was derived from a survey item (“Where in your home do you store your prescriptions that you take on a regular basis?”) to which respondents could select all that applied: “Kitchen table,” “Kitchen cabinet,” “Kitchen counter,” “Kitchen drawer,” “In the refrigerator,” “On the bathroom vanity,” “In the vanity drawer or cabinet,” “Bathroom medicine cabinet,” “On top of the bedroom nightstand,” “In the nightstand drawer,” “Desk,” “Dining room table,” “Backpack, purse, or bag,” “Closet,” and an open-text selection for unlisted locations. Open-text responses indicating use of already existing values were categorized as those values accordingly via consensus coding to promote interrater reliability.

#### 2.3.6 Receptivity to Storage Location Guidance

Receptivity to storage guidance was measured using a survey item (“If you received a new prescription, would you be open to receiving guidance on where to store the medication?”) to which respondents could select all that applied (“Yes, from my physician,” “Yes, from my pharmacist,” “Yes, on an app or website,” “Yes, in a brochure,” “No”). All “yes” responses were consolidated into a binary variable indicating guidance receptivity to one or more listed sources (physician, pharmacist, mobile app or website, or brochure). Responses of “No” were cleaned by excluding any response for whom a “yes” response was also given for the item.

### 2.4 Analysis

Relative frequencies were assessed for all sample characteristics and variables listed in our *Measures* subsection. Bivariate analyses were conducted for digital medication reminder method variables and variables indicating adherent behavior to medications taken regularly, receptivity to medication storage guidance, and medication storage locations currently in use by respondents. Analyses included chi^2^ tests of homogeneity of proportions and bivariate logistic regression models.

## 3 Results

We report on the results of our analysis of survey responses.

### 3.1 Sample Characteristics

Respondents in our older adult sample were 59% female and 40% male. Less than 1% of respondents identified with a non-listed sex. Respondents ranged from 40 to over 90 years old with most (64%) between the ages of 40 and 59 years. Most respondents self-identified as white (76%). Of white respondents (n=439), the vast majority (95%) selected no other race or ethnicity. Just over 12% of the sample identified as American Indian or Alaskan Native, 7% identified as Black or African American, 5% identified as Asian, 5% identified as Hispanic or Latino, 2% identified as Native Hawaiian or Pacific Islander, and less than 1% identified with an unlisted race or ethnicity. Respondents’ education levels varied, though most respondents (52%) identified as having obtained a bachelor’s or master’s degree (Table 1).

### 3.2 Digital and Non-digital Medication Reminders

Over half of respondents (56%) indicated that they use digital methods to remember to take their medication and 65% indicated that they use non-digital methods. The most common digital methods reported included use of smartphone applications or alarms (39%). The most common non-digital method reported was use of labeled pillboxes (36%). More than half of respondents (54%) reported using a combination of both digital and non-digital methods to remember to take their medication.

### 3.3 Perceived Importance of Medication Adherence and Adherent Behavior

Most respondents (89%) indicated that they felt taking medication as prescribed was “important” or “very important,” and 66% had not forgotten to take a medication in the two weeks prior to survey. Over 71%, however, indicated that they had ever forgotten to take a medication. Of those who had ever forgotten to take a medication (n=415), almost half (48%) had forgotten to take a medication in the two weeks prior to responding to the survey.

### 3.4 Home Medication Storage Locations

The most popular home medication storage locations for medications taken regularly by our sample included nightstand drawers (27%), kitchen cabinets (25%), and atop bedroom nightstands (23%). Despite the name, only 19% of our sample stored medication in medicine cabinets. Other locations included kitchen counters (16%), in refrigerators (14%), in vanity drawers or cabinets (14%), inside of bathroom vanities (14%), in kitchen drawers (14%), inside or on top of desks (14%), in backpacks, purses, or bags (11%), on kitchen tables (10%), on dining room tables (8%), and in closets (4%). Less than 3% of respondents stored medications principally in unlisted locations.

### 3.5 Receptivity to Storage Guidance

Most respondents (94%) indicated that they would be receptive to guidance about where to store their medications, with more preferring guidance from pharmacists (66%) followed by guidance from physicians (55%). Fewer (19%) indicated that they would be receptive to guidance delivered on a digital platform, such as websites or mobile applications.

### 3.6 Bivariate Analyses

#### 3.6.1 Use of Digital Reminder Methods and Medication Adherence

Bivariate analyses revealed statistically significant associations between use of any digital medication reminder methods (composite variable) and having ever forgotten to take a medication (p<0.001, chi^2^). Affirmative indication of having used any digital method was significantly associated with decreased odds of having ever forgotten to take a medication (unadjusted OR: 0.24, 95% CI: 0.16 – 0.36). Significant associations were not observed between use of digital methods (composite variable) and forgetting to take a medication in the two weeks prior to survey.

Digital medication reminder methods significantly associated with decreased odds of having ever forgotten to take a medication included use of smartphone applications and alarms (unadjusted OR: 0.49, 95% CI: 0.34 - 0.71), Apple Siri (unadjusted OR: 0.26, 95% CI: 0.16 - 0.42), smartwatches (unadjusted OR: 0.52, 95% CI: 0.32 - 0.84), and electronic pill dispensers (unadjusted OR: 0.49, 95% CI: 0.29 - 0.81). Use of other listed digital methods (Amazon Alexa and electronic attachments to pill bottles) was not significantly associated with having ever forgotten to take a medication.

Interestingly, while no digital reminder methods were significantly associated with decreased odds of having forgotten to take a medication in the two weeks prior to survey, some digital methods were associated with increased odds, including Amazon Alexa (unadjusted OR: 2, 95% CI: 1.07 – 3.77) and electronic pill dispensers (unadjusted OR: 1.68, 95% CI: 1.02 – 2.76).

#### 3.6.2 Use of Digital Reminder Methods and Receptivity to Storage Guidance

Use of any digital medication reminder methods (composite variable) was significantly associated with increased odds of receptivity to guidance from one or more listed sources (composite variable), including from physicians, pharmacists, mobile applications or websites, or brochures (unadjusted OR: 3.99, 95% CI: 1.83 – 8.67). However, associations between use of any digital reminder methods and preferred source of medication storage guidance were only significant for the responses of “pharmacists” (unadjusted OR: 0.63, 95% CI: 0.44 – 0.9) and “brochures” (unadjusted OR: 0.32, 95% CI: 0.21 – 0.48), both of which indicate that use of digital methods was associated with decreased odds of receptivity to guidance from this source.

#### 3.6.3 Use of Digital Reminder Methods and Medication Storage Locations

Bivariate analyses indicated significant associations between the use of any digital medication reminder methods (composite variable) and increased odds of using certain home medication storage locations, including kitchen tables (unadjusted OR: 2.22, 95% CI: 1.22 – 4.04), kitchen counters (unadjusted OR: 1.70, 95% CI: 1.06 – 2.72), kitchen drawers (unadjusted OR: 5.98, 95% CI: 3.09 – 11.57), refrigerators (unadjusted OR: 3.5, 95% CI: 1.99 – 6.14), vanity drawers or cabinets (unadjusted OR: 1.92, 95% CI: 1.16 – 3.17), atop bedroom nightstands (unadjusted OR: 2.93, 95% CI: 1.91 – 4.51), in nightstand drawers (unadjusted OR: 2.49, 95% CI: 1.68 – 3.69), inside or on top of desks (unadjusted OR: 6.73, 95% CI: 3.39 – 13.35), dining room tables (unadjusted OR: 3.51, 95% CI: 1.66 – 7.42), and backpacks, purses, or bags (unadjusted OR: 3.13, 95% CI: 1.69 – 5.80).

#### 3.6.4 Medication Storage Locations and Medication Adherence

Several home medication storage locations were significantly associated with decreased odds of having ever forgotten to intake a medication, including kitchen drawers (unadjusted OR: 0.51, 95% CI: 0.31 – 0.83), in refrigerators (unadjusted OR: 0.57, 95% CI: 0.35 – 0.93), atop bedroom nightstands (unadjusted OR: 0.52, 95% CI: 0.35 – 0.78), in nightstand drawers (unadjusted OR: 0.38, 95% CI: 0.26 – 0.56), desks (unadjusted OR: 0.33, 95% CI: 0.21 – 0.54), and backpacks, purses, or bags (unadjusted OR: 0.5, 95% CI: 0.29 – 0.85). Two home medication storage locations were significantly associated with increased odds of having ever forgotten to intake a medication: kitchen cabinets (unadjusted OR: 1.57, 95% CI: 1 – 2.45) and bathroom vanities (unadjusted OR: 4.12, 95% CI: 1.94 – 8.76). All remaining home medication storage locations were not associated with having ever forgotten to intake a medication.

Far fewer home medication storage locations were significantly associated with having forgotten to take a medication in the two weeks prior to survey alone. Only one location was significantly associated with decreased odds of forgetting to intake a medication in the two weeks prior to survey: nightstand drawers (unadjusted OR: 0.56, 95% CI: 0.37 – 0.84). Two locations were significantly associated with increased odds of forgetting to intake a medication in the two weeks prior to survey: kitchen tables (unadjusted OR: 1.9, 95% CI: 1.09 – 3.28) and kitchen cabinets (unadjusted OR: 1.71, 95% CI: 1.16 – 2.51).

## 4 Discussion

### 4.1 Importance of and Self-reported Adherence

Most respondents (89%) indicated that they felt that taking medication as prescribed was “important” or “very important,” indicating that they may be more likely to be receptive to medication adherence interventions. The responses for the two questions on whether the respondent had forgotten to take a medication in the past two weeks or ever were used to measure medication nonadherence (34). Most (66%) had remembered to take all medications in the two weeks prior to survey. Over 70%, however, indicated that they had forgotten to take a medication at least once in their lives. We need to further explore how these findings relate to other studies that have explored both medication adherence and nonadherence (35).

### 4.2 Home Medication Storage Locations

Despite the name “medicine cabinet,” we found that the most commonly used storage locations for medications taken regularly by our sample were nightstand drawers (27%), kitchen cabinets (25%), and atop bedroom nightstands (23%). The survey results confirmed our hypothesis that patients choose a variety of locations to store their medications, and we next will learn more about the reasons underlying these choices. Some reasons may be related to home conditions, since two of the most popular locations are likely to be personal no matter the number of people in the home. Our study did not isolate locations that were selected for climactic requirements or safety, which may also be factors.

The University of Michigan National Poll on Healthy Aging is the only previous study that considered storage location, although their study interest was grandchild safety (24). The University of Michigan National Poll was completed by 1,074 adults between the ages of 50-80 with a grandchild 0-17 years of age, while the subset of our study analyzed here was completed by 580 respondents 40 years of age and older. In the two studies, respondents were given different lists of location options to choose from and the naming was also different for some of the locations (e.g., “kitchen cabinet” compared to “cupboard or cabinet”). The locations reported from the Tufts University School of Medicine and the University of Michigan National Poll on Healthy Aging surveys differ greatly, which may be attributed to the options available to respondents. Neither survey asked about the characteristics of the locations nor the determinants of location selection.

Our exploratory analyses suggest significant bivariate associations between several home medication storage locations and decreased odds of having ever forgotten to intake a medication, including atop nightstands and inside nightstand drawers. Other locations, including kitchen cabinets, were associated with increased odds of having ever forgotten to intake a medication. Fewer locations were associated with decreased odds of having forgotten to intake a medication in the two weeks prior to survey.

Since these unadjusted, exploratory analyses suggest that some home medication storage locations may be associated with adherence, the next step in our research is to learn what factors influence location selection, understand the implications for adherent behavior, and take apply more sophisticated multivariable approaches to our analysis.

Most survey respondents (94%) indicated that they would be receptive to guidance regarding where to store their medications, with more preferring guidance from a pharmacist (66%) followed by guidance from physicians (55%). In subsequent research, we hope to learn either if the characteristics of the locations or the determinants of location selection are associated with adherence. Knowing respondents’ receptivity to guidance, and knowing more about the selection of locations associated with adherence, in our next study we plan to design interventions to guide storage selection.

### 4.3 Medication Reminders

There are many devices and apps that offer reminders to take medication. Devices to increase adherence have not yet shown their effectiveness (31, 35, 36). There may be many reasons for this including the lack of incorporation of location as well as the focus on time-based notifications.

No known devices to dispense and/or remind include recommendations for optimal device storage locations, yet, since many devices use auditory and/or visual cues, we suspect that location may be a consideration for cues to be heard or seen. Further, no known devices are designed for specific locations in the home. Over half of respondents (56%) indicated that they use digital methods to remember to take their medication, the most common being the use of smartphone applications or alarms (39%). Over 65% of respondents indicated that they use non-digital methods, of which the most common was use of labeled pillboxes (36%). More than half of respondents (54%) reported using both digital and non-digital methods to remember to take their medication. The sheer number of respondents using reminders is indicative of a perceived need, yet the lack of effectiveness in studies is concerning.

Our analysis found no significant associations between use of digital methods and forgetting to take a medication in the two weeks prior to survey. However, digital medication reminder methods that were significantly associated with decreased odds of having ever forgotten to take a medication included use of smartphone applications and alarms, Apple Siri, smartwatches, and electronic pill dispensers. Interestingly, while no digital reminder methods were significantly associated with decreased odds of having forgotten to take a medication in the two weeks prior to survey, some digital methods were associated with increased odds, including Amazon Alexa and electronic pill dispensers. Furthermore, the use of any digital medication reminder method was significantly associated with increased odds of receptivity to guidance from one or more of the sources listed, including from physicians or pharmacists.

In subsequent analyses, we will examine other survey responses. One question asked about the frequency with which respondents check for unused or expired medications, which may be a proxy for conscientious medication management. Mere reduction of clutter in medication storage may increase adherence with fewer prescription bottles to choose from. Since sudden changes in lifestyle can influence medication adherence, we asked a question about changes in adherence in the Coronavirus (COVID-19) pandemic to learn if more time at home eased medication management and thus increased adherence; alternatively, the changes in routine (including increased rates of pharmacy pick-up) may have reduced adherence.

Finally, in planned qualitative studies we hope to better understand the selection of medication storage locations, the use of rooms in the home, and how the spaces within those rooms relate to routines and cues to assist in those routines (28). Through these interviews, we hope to understand the relationship between medication adherence and factors we have not previously explored, one being patient activation, which comprises the degree of knowledge, confidence, and skills that patients have to manage their overall health (37).

## 5 Limitations

As our survey recruitment was primarily achieved via a mailing list and social media, our sample is not representative of the US population. Additionally, our exploratory analyses were bivariate and thus did not adjust for other additional variables.

## 6 Conclusions

Our long-term goal is to aid older adults in making informed choices about where to store their medications in their homes to increase adherence and to design interventions that support them in implementing these choices in their homes. Strong evidence to guide optimization of storage locations could play a crucial role in improving adherence and, thereby, the health and safety of older adults living at home. Based on survey results, we are encouraged to develop best practice techniques to counsel older adults on optimal selection of home medication storage locations to improve their medication adherence, delivered by pharmacists and physicians. Our future research will investigate if there is a relationship between medication storage locations and adherence by more fully understanding the characteristics of storage locations and the determinants of location selection. By understanding which factors related to storage location impact adherence, we hope to design innovative digital solutions that promote better medication habits. Our future research will also investigate how new device designs can incorporate specific storage locations and contextual cues related to location to more effectively promote adherence. With a rising number of older adults and a commensurate increase in the number of patients taking prescription medications, interventions to increase adherence will lead to greater health and longevity.

## Data Availability

All data produced in the present study are available upon reasonable request to the authors and subject to agreement by the Tufts University health Sciences IRB.

## 8 Author Contributions

The authors are responsible for developing and deploying the survey and all analysis of data reported here. LG initiated this research project, was responsible for developing and deploying the survey, and contributed to this paper. ES performed the primary analysis of the survey and contributed to this paper. BE and AV contributed to this paper.

## 10 Acknowledgments

The authors offer their appreciation to the graduate students at Tufts University School of Medicine who contributed to this work, in particular, Justin Barton, Cheryl Croll, Meera Singhal, and Ricardo Boschetti.

## 11 Supplementary Material

